# Development of Goal Management Training^+^ (GMT^+^) for Methamphetamine Use Disorder Through Collaborative Design: A Process Description

**DOI:** 10.1101/2021.08.01.21261454

**Authors:** Alexandra C. Anderson, Alex H. Robinson, Eden Potter, Bronte Kerley, Daphne Flynn, Dan I. Lubman, Antonio Verdejo-García

## Abstract

**Background:** Methamphetamine use disorder (MUD) is associated with executive dysfunctions, which are linked with poorer treatment outcomes including earlier drop out and relapse. However, current treatments for MUD do not address executive functions. Goal Management Training (GMT) is an evidence-based cognitive remediation program for executive dysfunction but required modifications to enhance its relevance and application within addiction treatment settings. This study aimed to 1) tailor GMT to the key cognitive deficits and typical treatment duration of MUD; 2) explore consumers’ (people with MUD) engagement with the revised program; 3) implement a protocol of the program with consumers; and 4) present the manualised standard administration to clinical service providers.

**Methods:** We followed the Medical Research Council Complex Interventions Framework and employed an evidence- and person-based intervention development process. We used a four-phased approach and collaborated with neuropsychology experts, design researchers in healthcare, consumers with MUD, and clinical service providers. Each aim was addressed in a separate study phase; including content refinement and review with neuropsychology experts (phase 1), intervention design and collaboration with consumers (phase 2), prototype development and review with consumers (phase 3), and final program modifications and review with clinical stakeholders (phase 4).

**Results:** Findings from phase 1 indicated support for targeting four cognitive processes (attention, impulse control, goal setting, and decision-making). Key feedback included the need to help habitualise program strategies and guide consumers in applying them in emotionally salient situations. Findings from phases 2 and 3 indicated consumer support for the program strategies and materials but highlighted the need to further enhance the personal relevance of specific content and journal activities. Findings from phase 4 provided overall clinical support for the revised program but indicated an opportunity to minimise unintended effects. We present the intervention materials for the revised program, Goal Management Training^+^ (GMT^+^), in line with TIDieR guidelines.

**Conclusions:** GMT^+^ targets tailored cognitive processes, includes reimagined materials and activities, and is sensitive to the clinical needs of people with MUD. Our intervention development process was important for informing the training components, design and intervention materials, and indicating initial acceptability prior to conducting a clinical trial.

## Background

Methamphetamine is a highly addictive stimulant that presents a global public health concern^1^. In 2019, approximately 27 million people had used amphetamines worldwide, and there is growing concern around the rise of harmful patterns of use^2^. Methamphetamine use disorder (MUD) is associated with greater risk of suffering physical and mental health conditions, including cardiovascular disease, blood-borne viruses, psychosis, depression, and suicide, as well as social disadvantage^3,4,5^. Underlying the hallmark characteristics of MUD (i.e., loss of control over drug intake, escalation of use despite growing negative consequences) are cognitive deficits in executive functions (the higher-order cognitive skills that orchestrate goal-directed behaviours)^6, 7^.

Emerging research has revealed that executive functions, such as inhibitory control, working memory and decision-making, are significantly associated with MUD treatment outcomes^8, 9^. Specifically, consumers with deficits in executive functions are at greater risk of dropping out of treatment, relapsing after abstinence-oriented treatment, and struggling to regain quality of life^10, 11^. This research, together with recent evidence showing that current treatment interventions for MUD have overall limited efficacy^4^, raises the need to incorporate cognitive remediation interventions for executive dysfunction as an add-on to current treatment approaches^12, 13^. Cognitive remediation interventions are targeted to strengthen executive functions via meta-cognitive skills and strategy learning within a therapeutic group environment^14^.

In a recent meta-analysis of cognitive-boosting interventions for addiction treatment, we showed that Goal Management Training (GMT) is the most promising approach to ameliorate executive deficits in this context^15^. GMT was originally developed to improve executive functions in brain injury populations^16^, but its active ingredients, such as mindfulness and strategies to prevent disinhibited responses and manage complex tasks, are well suited for substance use disorders^17,18,19,20^. However, the original GMT presents three key limitations in the context of MUD treatment. Firstly, the program is lengthy, i.e., 7 to 9 weeks, which almost doubles the standard duration of treatment episodes for MUD^21^. Second, the training activities and their delivery were not designed to address the nature and severity of cognitive deficits in substance use disorders or MUD specifically. In MUD, deficits are less pronounced than in brain injury, and there is a need for a greater emphasis on aiding long-term decision-making and inhibiting impulsive behaviours, which are key predictors of addiction treatment outcomes^8^. Third, the presentation of materials (including character examples, design, and activities) may lack engagement potential for people with MUD. The original program was designed to suit older adults with different demographics and may not be adequate to capture attention or personal relevance for people with MUD. In addressing these limitations, it is important to incorporate the views of people who use methamphetamine to enhance the intervention experience for the end-consumers^22^ and ultimately to increase the chance of it being considered as ‘helpful’ and acceptable.

The purpose of this study was to develop a modified version of Goal Management Training (now Goal Management Training^+^; GMT^+^) to strengthen executive function and improve clinical outcomes in individuals with MUD. The results of this study will inform a proof-of-concept trial of GMT^+^ for people with MUD. Our specific aims were:

**Aim 1**: To develop an updated version of GMT tailored to the cognitive deficits of people with MUD and the duration of typical treatment episodes for MUD / substance use disorders.

**Aim 2**: To gather consumers’ (i.e., people with MUD) engagement with the updated GMT program.

**Aim 3**: To implement a consumer-acceptable prototype of the program (i.e., GMT^+^).

**Aim 4:** To manualise the intervention, showcase a standard administration among clinicians, and prepare materials for the proof-of-concept pilot trial.

## Methods and Results

### Design

We followed the Medical Research Council Complex Interventions Framework^23^. The intervention development process was underpinned by an evidence- and person-based approach. This approach aims to ground the development of interventions in a deep understanding of the consumer perspective, with consideration to their psychosocial context^24^. Importantly, the process involves a flexible approach that is guided by understanding the needs, goals, and desires of the end consumers (i.e., people with MUD)^25^. This is activated through involving consumers in the development process as experts of their own life experiences, within a participatory design framework^26^. The intervention development process incorporates multidisciplinary skills and perspectives by including clinical psychology researchers (core research team), neuropsychology researchers, design researchers in healthcare, consumers with MUD, and clinical service providers.

We used a qualitative approach and developed the intervention in four phases. Phase 1 involved content refinement, where the existing content was assessed for relevance for MUD and reorganised in a streamlined set of modules / sessions to bring it closer to the standard duration of MUD treatment. We then conducted a focus group with experts in neuropsychology to evaluate these changes and seek further recommendations. This was considered the intervention planning phase. Phase 2 involved intervention design. We collaborated with design researchers in healthcare to reimagine the materials and visual identity for GMT^+^, as well as to increase the experiential engagement of activities involving the core GMT skills. Next, we conducted a focus group with consumers with MUD to gain feedback on the design and engagement with the key concepts and activities. Intervention development took place over phases 3 and 4. Phase 3 involved developing a prototype of the intervention. We conducted a second online focus group with people with MUD to review the changes from the first focus group and to test sample activities from the module and daily journal. Phase 4 included the final program modifications, including feedback gained in a review session with clinicians. We present a description of the final materials, in line with TIDieR guidelines^27^.

The Monash University Human Research Ethics Committee (12364) and Eastern Health Human Research Ethics Committee (LR19/023) approved the study and all participants provided informed consent. The full study, including the four phases, took place between March 2019 – November 2020.

### Phase 1: Planning – Abbreviating GMT

#### Aim

To develop an updated version of GMT that was tailored to the cognitive deficits of people with MUD, and to abbreviate the program to four weeks (in alignment with the typical duration of MUD treatment).

#### Methods

##### Preliminary work on re-development

The first step in re-developing GMT for MUD was to decide on the core training ingredients of the program, also with a view to streamline content to make it suitable for treatment services. We based our selection of key cognitive targets on meta-analyses and systematic reviews that we had recently conducted^8, 15, 28^. We also conducted brainstorming sessions within the core research team (AA, AR, AVG) to share recent experiences with relevant consumers, end-users and treatment services that could benefit from GMT. After deciding on the key elements to include in the updated program, we revised the contents and language to meet the needs of individuals with MUD. We then presented this proposal to a group of neuropsychology experts familiar with the original GMT program (focus group 1).

##### Focus group 1: neuropsychology researchers

We conducted a 2-hour focus group with neuropsychology researchers (n = 4). We sought feedback on the new program structure (i.e., abbreviated format and stronger focus on decision-making), whether the active GMT training ingredients were maintained and if the new elements were appropriate for the program, and whether the overall training ingredients were appropriate in the context of addiction. Facilitators (AA and AR) took written notes throughout the session to capture verbal feedback. After the focus group, the core research team met to review the data that were collected.

#### Results

##### Preliminary work on re-development

We prioritised a focus on enhancing attention, gaining control over impulsivity, and goal setting from the original GMT program. We also prioritised developing a long-term mindset. In response to this target, we strengthened the decision-making components of GMT, with a focus on long-term goals. One example of this is the incorporation of episodic future thinking (EFT) in module 4 (Decide). EFT involves imagining future events through guided instruction and aims to promote a longer-term mindset^29^. EFT has shown efficacy at improving preference for larger delayed rewards over smaller immediate rewards in substance-using populations^30, 31^. The language was updated to target a younger adult population (previously aimed at older adults) and is more aligned with therapeutic and conversational dialogue, rather than educational training language.

We proposed to restructure GMT into four 90-minute weekly modules. Each module was designed to train a specific cognitive function. Module 1 (Be Aware) trains focused attention, module 2 (Pause) trains impulse control, module 3 (Envision Goals) trains goal setting, and module 4 (Decide) trains decision-making. A breakdown of content in the original weekly program (GMT) and the revised 4-week program (GMT^+^) is presented in Table 1. The future-focussed decision-making strategies in module 4 (Decide), were not included in the original program. GMT^+^ is designed as a 4-staged cycle (see Figure 1) that can be employed in any moment, so that participants can be aware of their attention and surroundings, pause and breathe, consider their goals (short-term or long-term) and make a decision. We removed multiple variations of the cycle (e.g., STOP-State-Split; Stop-State-Split-Check) to avoid confusion and maintain a simplified message for the end-consumer. Concepts that were reinforced multiple times in the original program were reduced, although a brief recap is included at the start of each session.

**Figure 1.**
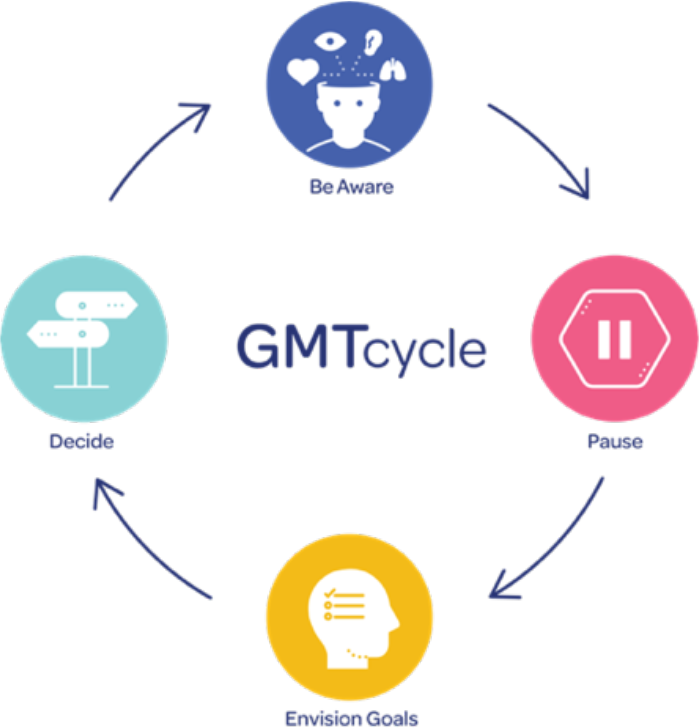
GMT^+^ cycle

**Table 1.**
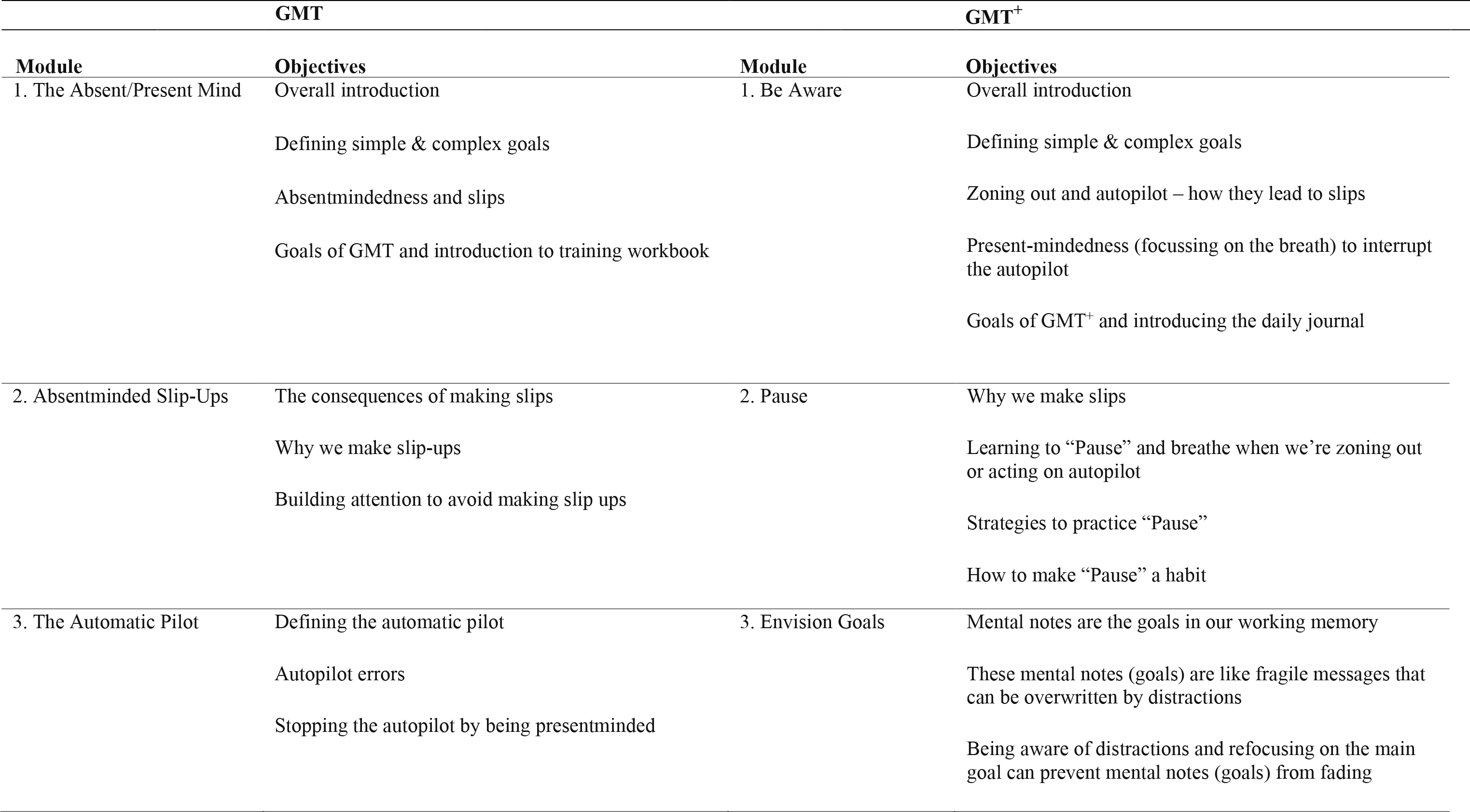
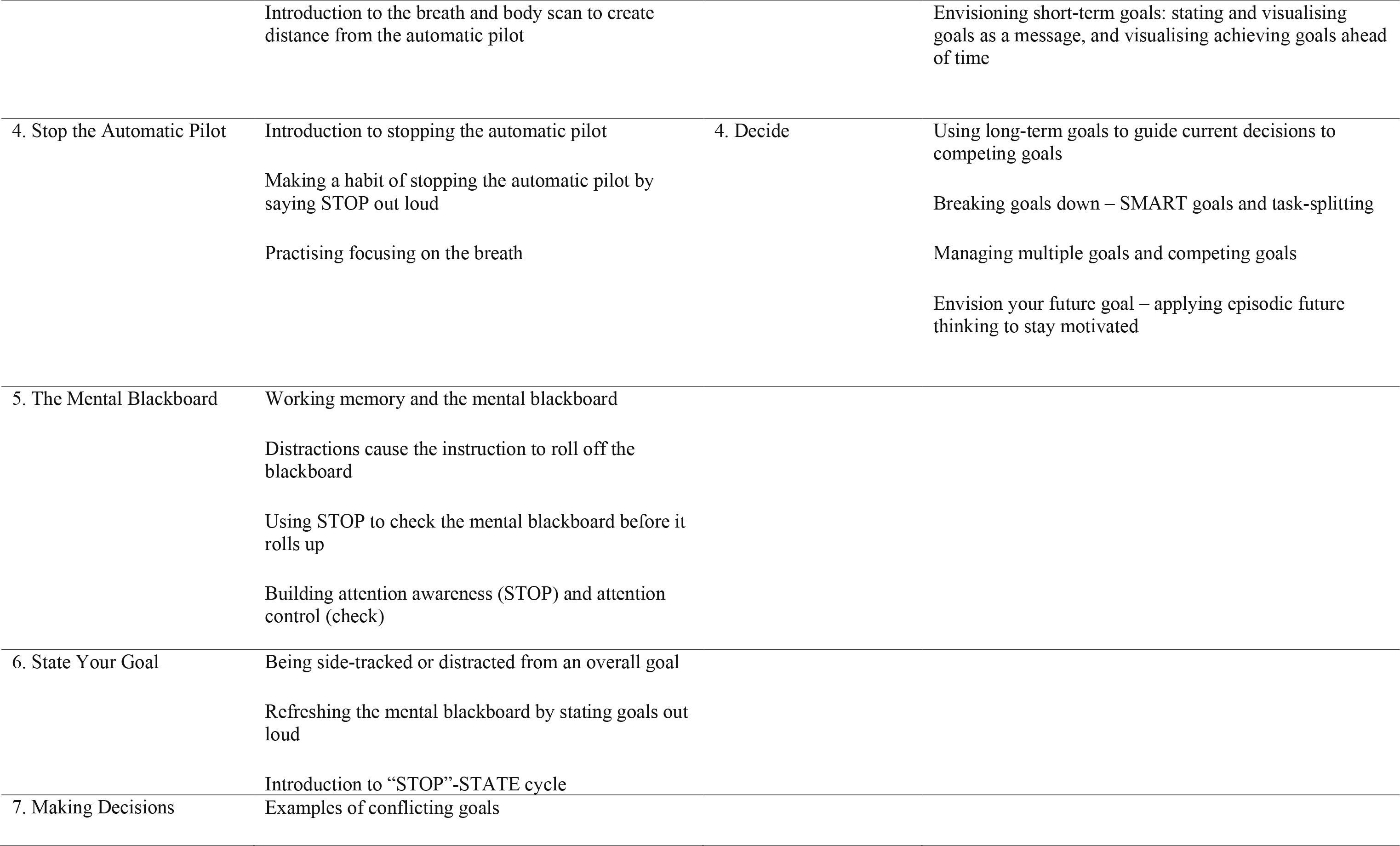
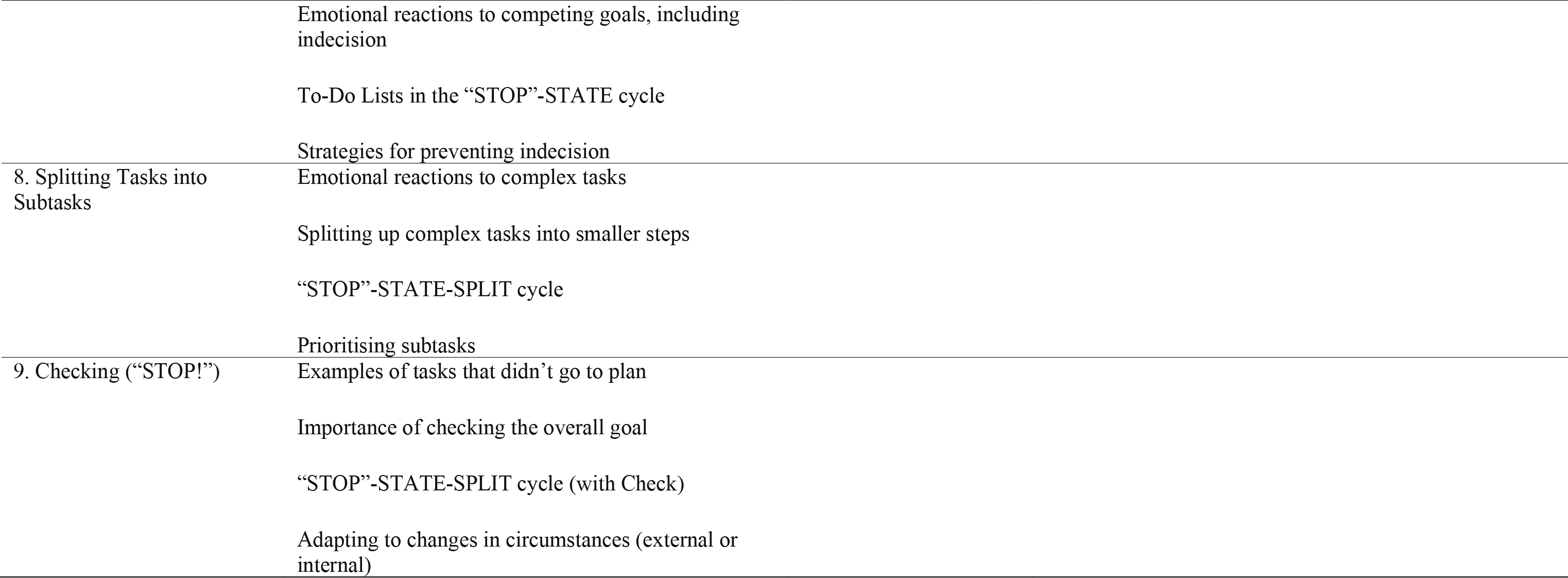
Revised program structure for GMT^+^

We selected in-session cognitive activities from the original program that matched GMT^+^ training principles, such as a simple routine-based task that elicits errors due to inattention (Be Aware module), and a multitasking activity that highlights goal neglect due to distractions (Envision Goals module). The activities in module 4 (Decide) were not designed to foster slips, but rather to help to participants to practice setting and staying on track with long-term goals (e.g., creating SMART goals).

##### Focus Group 1: neuropsychology researchers

Overall, neuropsychology experts were positive about the revised 4-session program structure. They further helped to select key learning activities (e.g., participant involvement in helping GMT characters to solve a problem) and approaches to delivering the content (e.g., including a mixture of theory, discussions, and practical activities). Key feedback from the session highlighted the importance of helping participants to habitualise the training concepts and to guide participants around how specific strategies (e.g., Pause) could be used in “hot” (i.e., in high level of emotionality) contexts (see Table 2). We responded to this feedback by prioritising the between-session journal engagement as a tool for skill habituation and agreed on the need to develop a visual reminder to employ strategies in everyday situations.

**Table 2.** Key feedback from the neuropsychology focus group and subsequent changes to the intervention planning.

| Theme | Feedback | Program modifications |
| --- | --- | --- |
| **New program content (abbreviated format and greater decision-making focus)** |  |  |
| Integrating theory with practice | Link the theory back to everyday behaviours that lead to drug-taking problems.<br>Minimise theory delivered in-session and maximise on ways of practising GMT<sup>+</sup> strategies.<br>Participants will need buy-in to the program, rather than too much theory. Explain the benefits at the beginning of the first session and consider talking about success stories. | - Included summary of the program's success in previous trials with addiction to promote buy-in at the start of session 1.<br>- Provided top-level theoretical information and then raised discussions about how this may relate to the group members.<br>- Increased examples of how and when GMT<sup>+</sup> skills can be practiced in everyday life. |
| Future goals need to be achievable | It is important to focus on achievable goals for participants. Reduce the possibility that they might set themselves up for failure with unachievable goals. | - Included exercises to help participants to think about goals that are achievable within 1 day, 1 month, 3 months, and 6 months.<br>- Increased in-session facilitator involvement to help participants to set realistic goals. |
| Anchor Pause to the Breath | Anchor the Pause command to relaxation and mindfulness.<br>One way to help generalise the skills to everyday life is to anchor them to the breath. | - Increased repetition of Pause-Breathe as a training concept in-session.<br>- Prioritised focussing on 'mindfulness' in the design phase to encourage regular focus on the breath. |
| **Appropriateness of new training ingredients in the context of GMT** |  |  |
| Testing new tasks and task variations | Experts suggested testing the task revisions to see whether they are too complex for people with MUD. | - Placed a priority on consumer testing of cognitive activities in the design phase.<br>- Assess whether they foster desired errors, or if they are perceived as too difficult. |
| Focus on task-splitting for decision-making | Task-splitting [from the original program] is important, particularly when participants are faced with rewarding decisions. | - Increased focus on task-splitting complex goals in module 4 (Decide). |
| **Relevance for addiction** |  |  |
| Reduce focus on past experiences | Avoid suggesting that participants look to the past to be reinforced by past behaviours and avoid talking about the past too much in group discussions [due to negative past experiences].<br>People with substance use disorders who have negative experiences may be highly sensitive to what hasn't worked. | - Reduced decision-making based on past consequences.<br>- Increased focus on future-based guided imagery experiences to visualise achieving a long-term goal (future-based motivation). |
| Enhancing relevance of language | Suggestion to update the language of training concepts.<br>For example, using everyday phrases like 'drifting thoughts', rather than 'absentmindedness'. | - Compiled different names for GMT concepts to test in the design phase.<br>- Language was placed as a priority for assessing acceptability. |
| The importance of Pause for MUD | This was considered as the most important concept by experts.<br>It will be important to help participants to habituate this concept.<br>Regular attendance and skills practice will be key to skill habituation. | - Provide in-session examples of when to "Pause" in everyday life.<br>- Encouraging attendance and between-session skills practice through engaging design. |
| Pause is important in the context of emotional valence | The program needs to help participants to focus on stopping thoughts about highly rewarding actions. | - Increased in-session content about Pausing when feeling triggered or stressed.<br>- Placed a priority on designing engaging skills practice activities to reinforce Pausing when feeling emotionally aroused. |

### Phase 2: Intervention Design – Enhancing Engagement

#### Aim

To gather consumers’ engagement with the updated GMT program. We conducted a focus group to assess initial acceptability of new additions to the program, aimed at enhancing consumers’ engagement. In terms of behavioural outcomes, we sought to increase the likelihood of regular attendance in sessions, as well as regular skills practice.

#### Methods

##### Preliminary work on re-imaging the materials

We collaborated with design researchers in healthcare to reimagine the intervention materials and enhance enjoyment of the program. Design priorities were grouped into five categories, including material re-design, program delivery, enhancing program relevance, assessing acceptability of program delivery, and encouraging skills practice. We prioritised ideas for a visual reminder to encourage participants to regularly practice GMT^+^ strategies (promoting skill habituation). Once the creative design phase was complete, we facilitated a focus group with individuals who were currently or previously seeking treatment for MUD, employing think-aloud techniques. We incorporated consumers’ perspectives early in the intervention development process to assess initial acceptability of the program, and prior to making final decisions on visual design elements and changes in cognitive tasks and journal activities

##### Focus Group 2: consumers with MUD (face to face session)

Focus group participants (n = 4) were recruited over a five-week period from Turning Point, a national addiction treatment centre based in Melbourne, or from eligible people who had previously participated in research with our group. Participants were compensated with a $20 grocery gift card. Eligibility criteria included a current or past diagnosis of MUD, currently seeking treatment or previous engagement in a treatment program for MUD, aged 18 or over, and the absence of intellectual disability or severe neurological condition. Seven participants consented to take part in the focus group, although three did not attend.

The focus group duration was two hours and was conducted at Turning Point. The format was a structured session with presentation slide content, guided questions, and ratings stickers to gauge preferences for select concepts. Participants were asked to share ideas, interact with tasks and materials, and provide verbal feedback. The facilitators assessed participants’ understanding by summarising key points and checking for accuracy. Facilitators took written notes throughout the session to capture verbal and non-verbal feedback. We sought feedback on material redesign (e.g., colour palettes, fonts, logo, illustrations), ways to enhance program relevance (discussing personal goals and previous treatment experiences), delivery format (how comfortable participants were in contributing to the group), and enhanced skills practice materials (presenting new activities, assessing difficulty level and whether they produced desired errors). The research team met after the focus group to review the data.

#### Results

##### Focus Group 2: consumers with MUD (face to face session)

Participants with MUD endorsed the novel four-staged cycle of Be-Aware-Pause-Envision Goals-Decide and considered GMT^+^ to be a valuable type of intervention that is currently missing from addiction treatment services. The results indicated initial acceptability of the creative journal activities, the GMT^+^ bracelet, and the group-based format that included discussions with and interaction between group members and facilitators. Interaction with the revised cognitive activities indicated that they appropriately elicited the desired errors to demonstrate executive dysfunction (e.g., missing specific details on the cartoon sorting cards). Participants thought the activities were aligned with the desired purpose and were enjoyable, with a minimum average enjoyment rating of 7/10 for each one. Qualitative themes and program changes are summarised in Table 3. Key feedback included examples of personal goals and the need for simplified creative activities. We used this information to facilitate relevant goal-related discussions and developed journal activities that permitted consumers to focus on working towards multiple goal categories throughout the program (e.g., short term and long term, or across different areas of life). We also prioritised the development of single-focussed creative activities that appeared relevant.

**Table 3.**
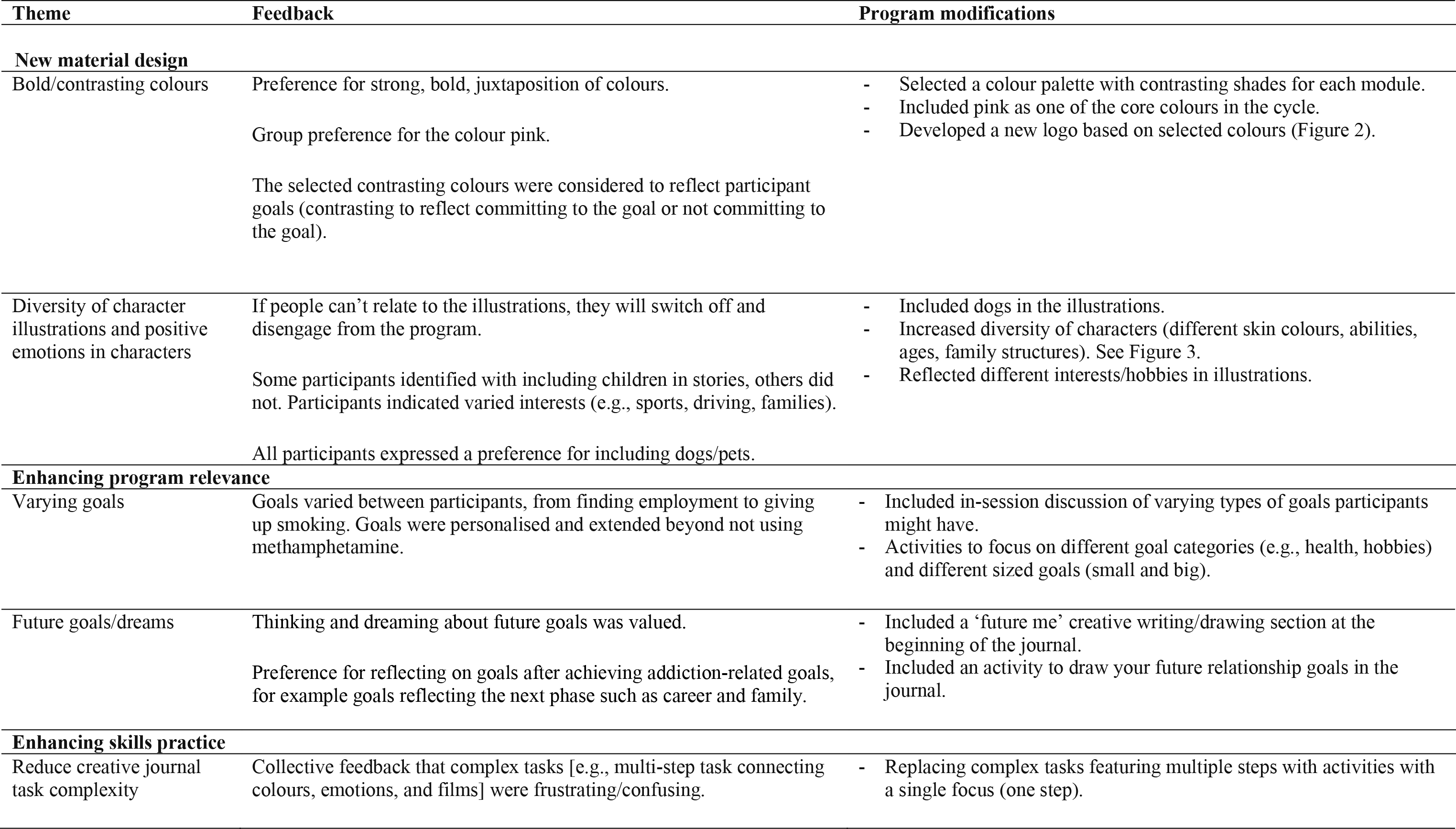
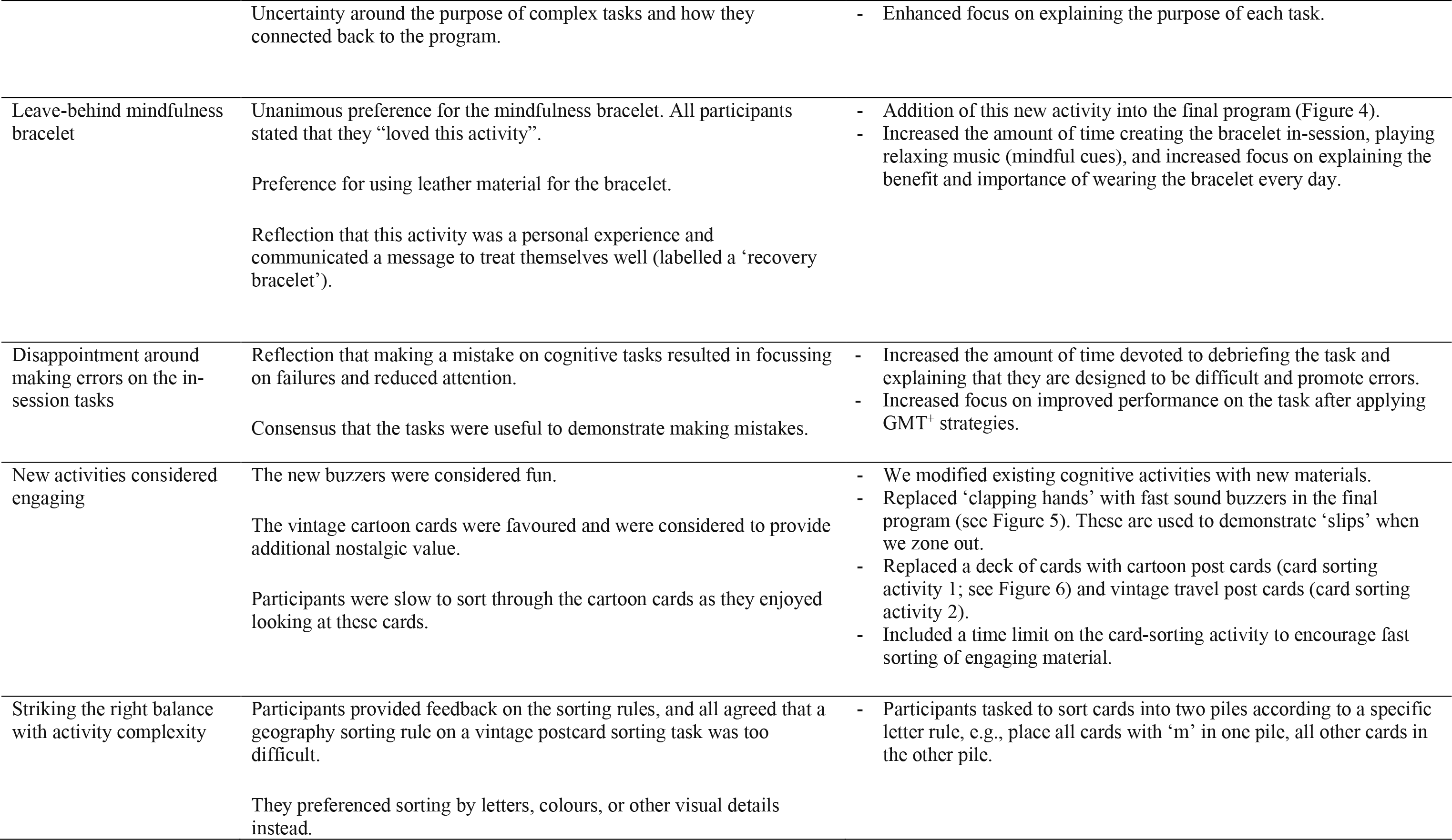
Key feedback from the design focus group with people with MUD and subsequent changes to the intervention design.

| Theme | Feedback | Program modifications |
| --- | --- | --- |
| **New material design** |  |  |
| Bold/contrasting colours | Preference for strong, bold, juxtaposition of colours.<br>Group preference for the colour pink.<br>The selected contrasting colours were considered to reflect participant goals (contrasting to reflect committing to the goal or not committing to the goal). | - Selected a colour palette with contrasting shades for each module.<br>- Included pink as one of the core colours in the cycle.<br>- Developed a new logo based on selected colours (Figure 2). |
| Diversity of character illustrations and positive emotions in characters | If people can't relate to the illustrations, they will switch off and disengage from the program.<br>Some participants identified with including children in stories, others did not. Participants indicated varied interests (e.g., sports, driving, families).<br>All participants expressed a preference for including dogs/pets. | - Included dogs in the illustrations.<br>- Increased diversity of characters (different skin colours, abilities, ages, family structures). See Figure 3.<br>- Reflected different interests/hobbies in illustrations. |
| **Enhancing program relevance** |  |  |
| Varying goals | Goals varied between participants, from finding employment to giving up smoking. Goals were personalised and extended beyond not using methamphetamine. | - Included in-session discussion of varying types of goals participants might have.<br>- Activities to focus on different goal categories (e.g., health, hobbies) and different sized goals (small and big). |
| Future goals/dreams | Thinking and dreaming about future goals was valued.<br>Preference for reflecting on goals after achieving addiction-related goals, for example goals reflecting the next phase such as career and family. | - Included a 'future me' creative writing/drawing section at the beginning of the journal.<br>- Included an activity to draw your future relationship goals in the journal. |
| **Enhancing skills practice** |  |  |
| Reduce creative journal task complexity | Collective feedback that complex tasks [e.g., multi-step task connecting colours, emotions, and films] were frustrating/confusing. | - Replacing complex tasks featuring multiple steps with activities with a single focus (one step). |
|  | Uncertainty around the purpose of complex tasks and how they connected back to the program. | - Enhanced focus on explaining the purpose of each task. |
| Leave-behind mindfulness bracelet | Unanimous preference for the mindfulness bracelet. All participants stated that they “loved this activity”.<br>Preference for using leather material for the bracelet.<br>Reflection that this activity was a personal experience and communicated a message to treat themselves well (labelled a ‘recovery bracelet’). | - Addition of this new activity into the final program (Figure 4).<br>- Increased the amount of time creating the bracelet in-session, playing relaxing music (mindful cues), and increased focus on explaining the benefit and importance of wearing the bracelet every day. |
| Disappointment around making errors on the in-session tasks | Reflection that making a mistake on cognitive tasks resulted in focussing on failures and reduced attention.<br>Consensus that the tasks were useful to demonstrate making mistakes. | - Increased the amount of time devoted to debriefing the task and explaining that they are designed to be difficult and promote errors.<br>- Increased focus on improved performance on the task after applying GMT<sup>+</sup> strategies. |
| New activities considered engaging | The new buzzers were considered fun.<br>The vintage cartoon cards were favoured and were considered to provide additional nostalgic value.<br>Participants were slow to sort through the cartoon cards as they enjoyed looking at these cards. | - We modified existing cognitive activities with new materials.<br>- Replaced ‘clapping hands’ with fast sound buzzers in the final program (see Figure 5). These are used to demonstrate ‘slips’ when we zone out.<br>- Replaced a deck of cards with cartoon post cards (card sorting activity 1; see Figure 6) and vintage travel post cards (card sorting activity 2).<br>- Included a time limit on the card-sorting activity to encourage fast sorting of engaging material. |
| Striking the right balance with activity complexity | Participants provided feedback on the sorting rules, and all agreed that a geography sorting rule on a vintage postcard sorting task was too difficult.<br>They preferred sorting by letters, colours, or other visual details instead. | - Participants tasked to sort cards into two piles according to a specific letter rule, e.g., place all cards with ‘m’ in one pile, all other cards in the other pile. |

**Figure 2.**
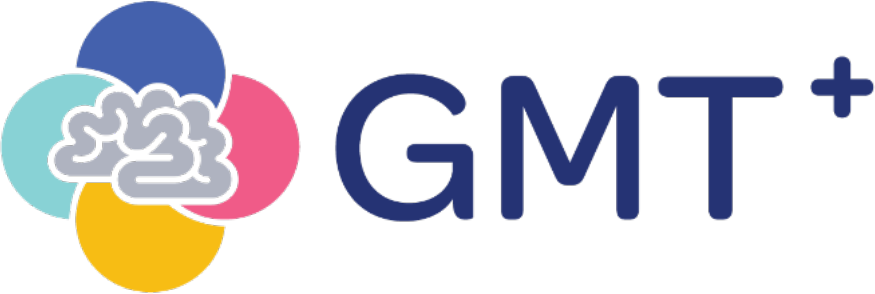
GMT^+^ logo. Each circle represents a module and a stage in the GMT cycle.

**Figure 3.**
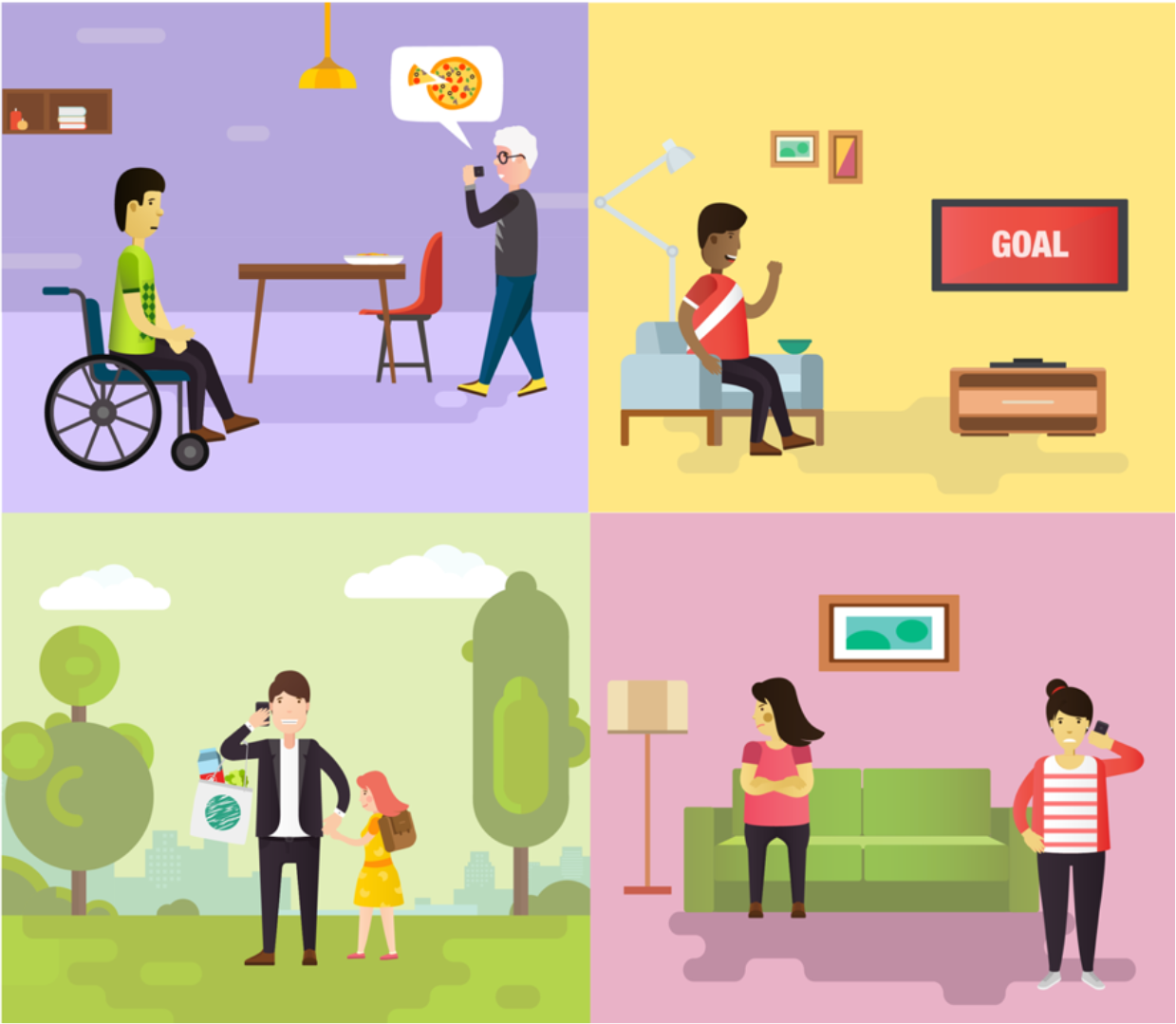
Examples of diverse GMT^+^ characters.

**Figure 4.**
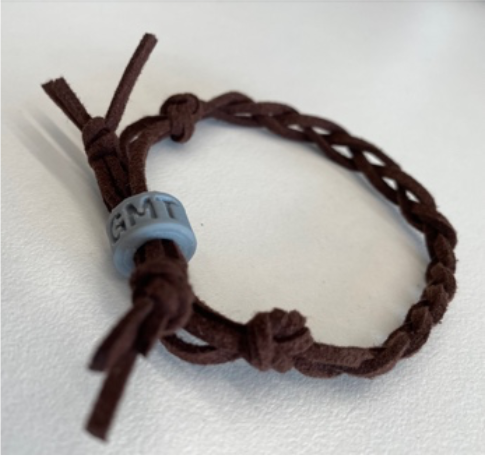
GMT^+^ mindfulness bracelet.

**Figure 5.**
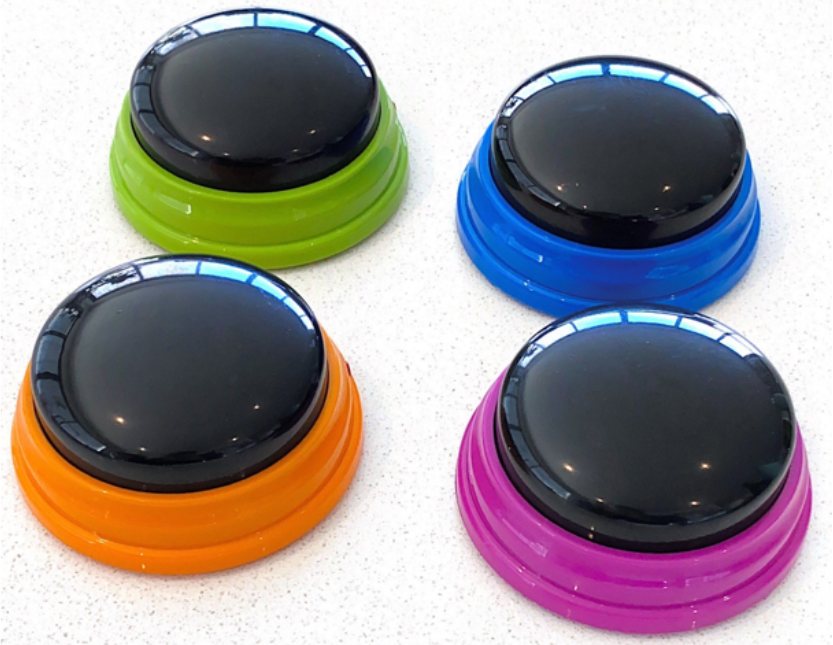
Interactive hand buzzers replaced a ‘hand-clapping’ task to enhance engagement.

**Figure 6.**
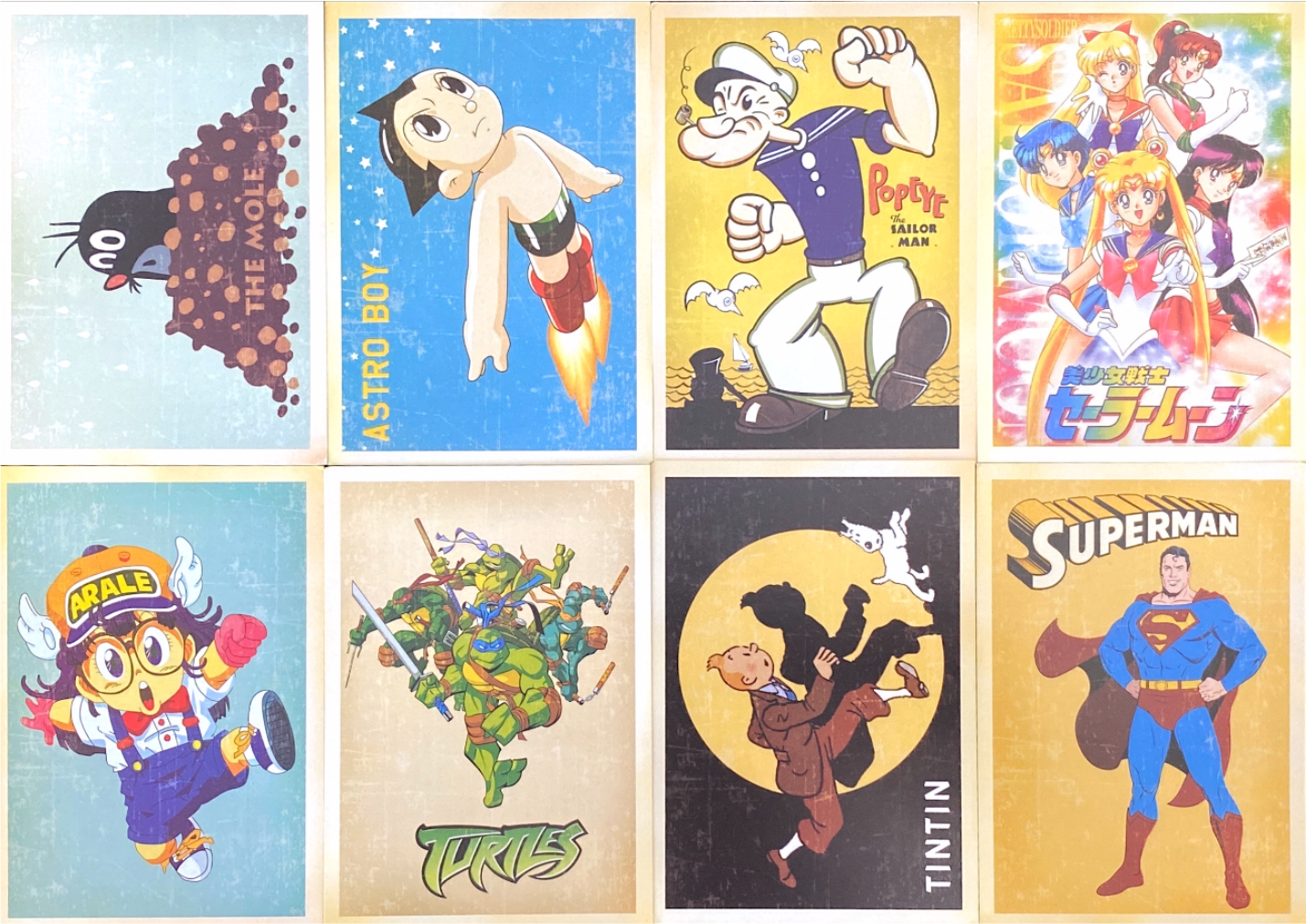
Example of new cartoon sorting cards.

### Phase 3: Intervention Development - Prototype

#### Aim

To develop and test a consumer-acceptable prototype of the updated program (GMT^+^). Specifically, we focussed on assessing consumer acceptability of GMT^+^ content, format, language, skills practice activities in the journal, and the delivery platform.

#### Method

##### Preliminary work on developing a prototype of the program

We collaborated with design researchers in healthcare and incorporated the feedback from phase 2 to develop a complete prototype of GMT^+^. The prototype included sample presentation slides and in-session activities relating to module 1 (Be Aware). We developed a printed journal with seven daily activities to encourage consumers to practice skills relating to module 1. This mindfulness-inspired journal includes both skills reflection activities (how participants used GMT^+^ skills in everyday life) and creative activities where they could practice GMT^+^ skills during the task (e.g., mindful drawing of the breath). Following development, we piloted a sample of the intervention using mixed methods. We conducted an online focus group with consumers with MUD. Qualitative aspects included observation of consumer engagement and their interaction style, and reviewing content themes that arose from think aloud techniques. Quantitative aspects included Likert scales to indicate acceptability of the journal content.

##### Focus group 3: consumers with MUD (teleconferencing session)

Five participants who were seeking treatment for MUD or had been recently engaged in treatment attended the online focus group. Three participants from focus group 2 (75%) returned for the follow-up online session, and we recruited two additional participants from Turning Point. Participants were compensated with a $25 grocery gift card for attending the focus group and an additional $15 grocery gift card for mailing back the completed journal sample and their feedback. Eligibility criteria was the same as focus group 2. In addition, participants needed to have access to the Internet and a device to access the video conferencing software (e.g., smart phone, tablet, laptop). Six participants consented to take part in the focus group and one participant did not attend on the day. The session was audio recorded and facilitators took written notes to capture non-verbal feedback.

The focus group duration was two hours and was conducted online, using videoconferencing software. The session included presentation slide content, guided questions, and ratings polls to gauge preferences. The prototype of module 1 was embedded in the session. We tested acceptability of the program and materials, the delivery format (including feasibility of an online delivery format), engagement with concepts and activities, engagement with the between-session journal, and the appropriateness of language.

Following the focus group session, we mailed out packs to attendees with the sample GMT^+^ journal and a pre-paid return envelope. Participants were asked to complete the journal daily, which was designed to take approximately one hour in total. A summary of the key training concepts was included in this journal. Participants were asked to time themselves completing each activity, to rate their engagement (Likert scales) and provide written feedback. After completing the journal, participants were asked to mail it back to the researchers. Qualitative feedback from the session and returned journals is included in Table 4.

**Table 4.**
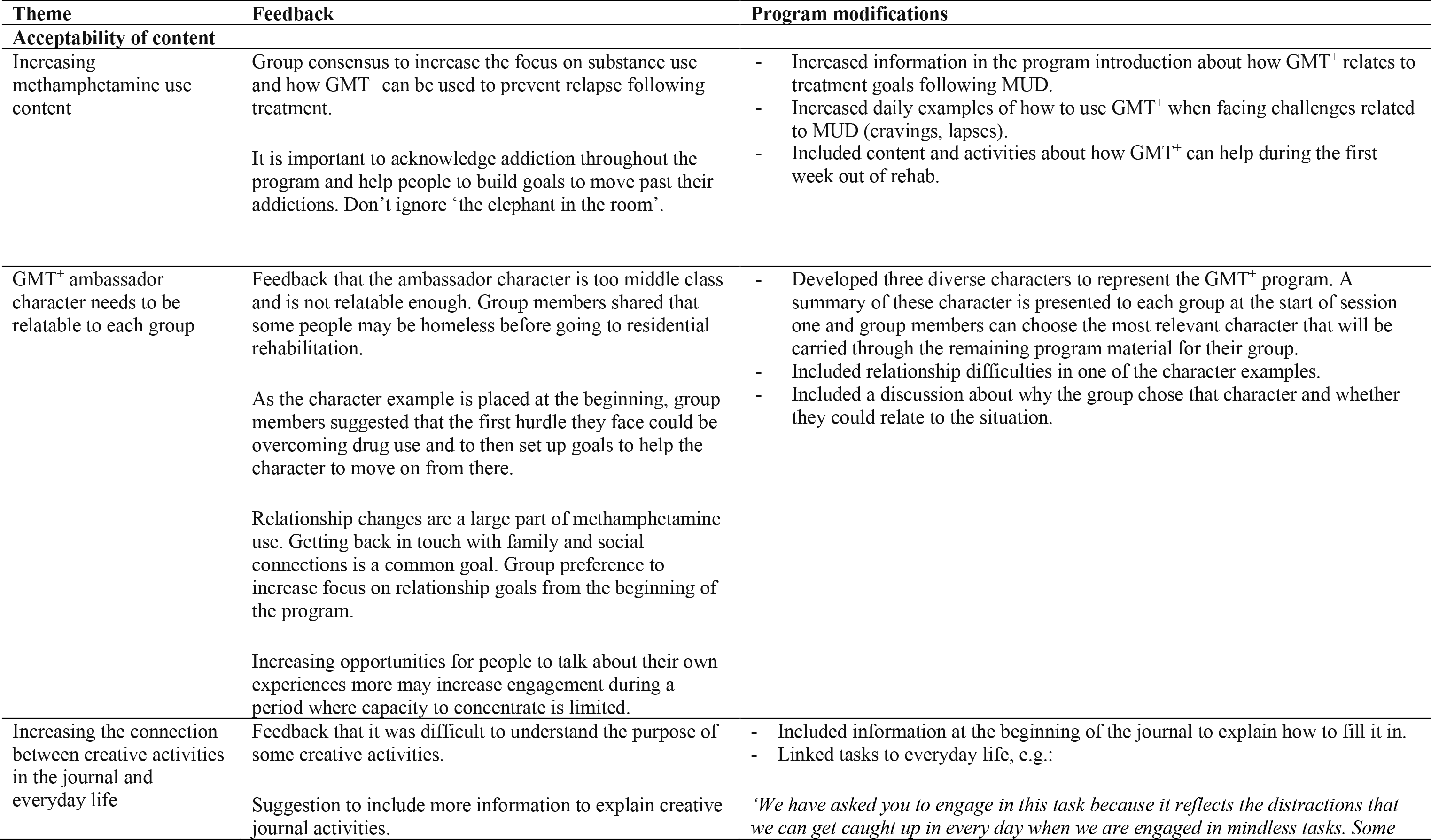
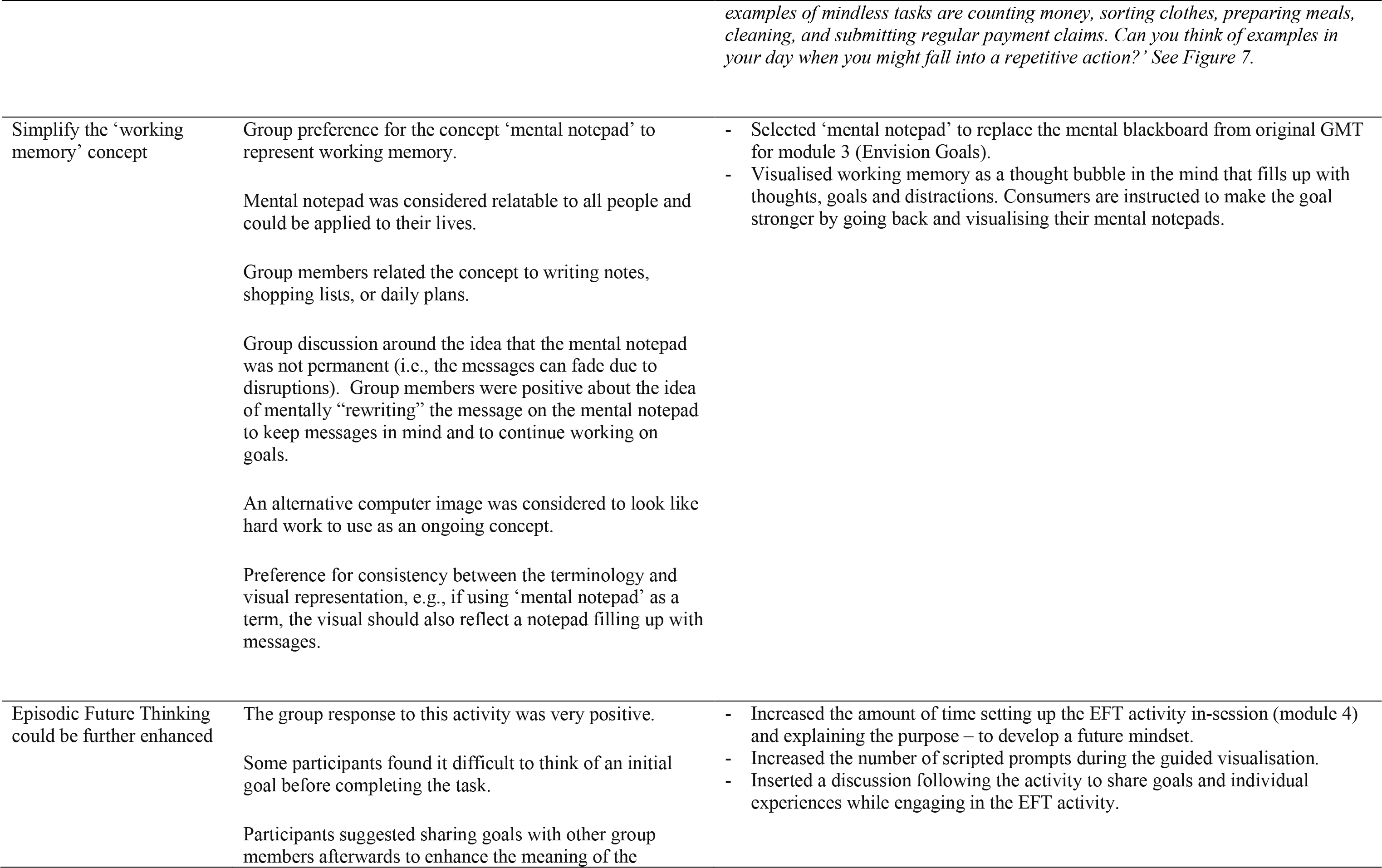
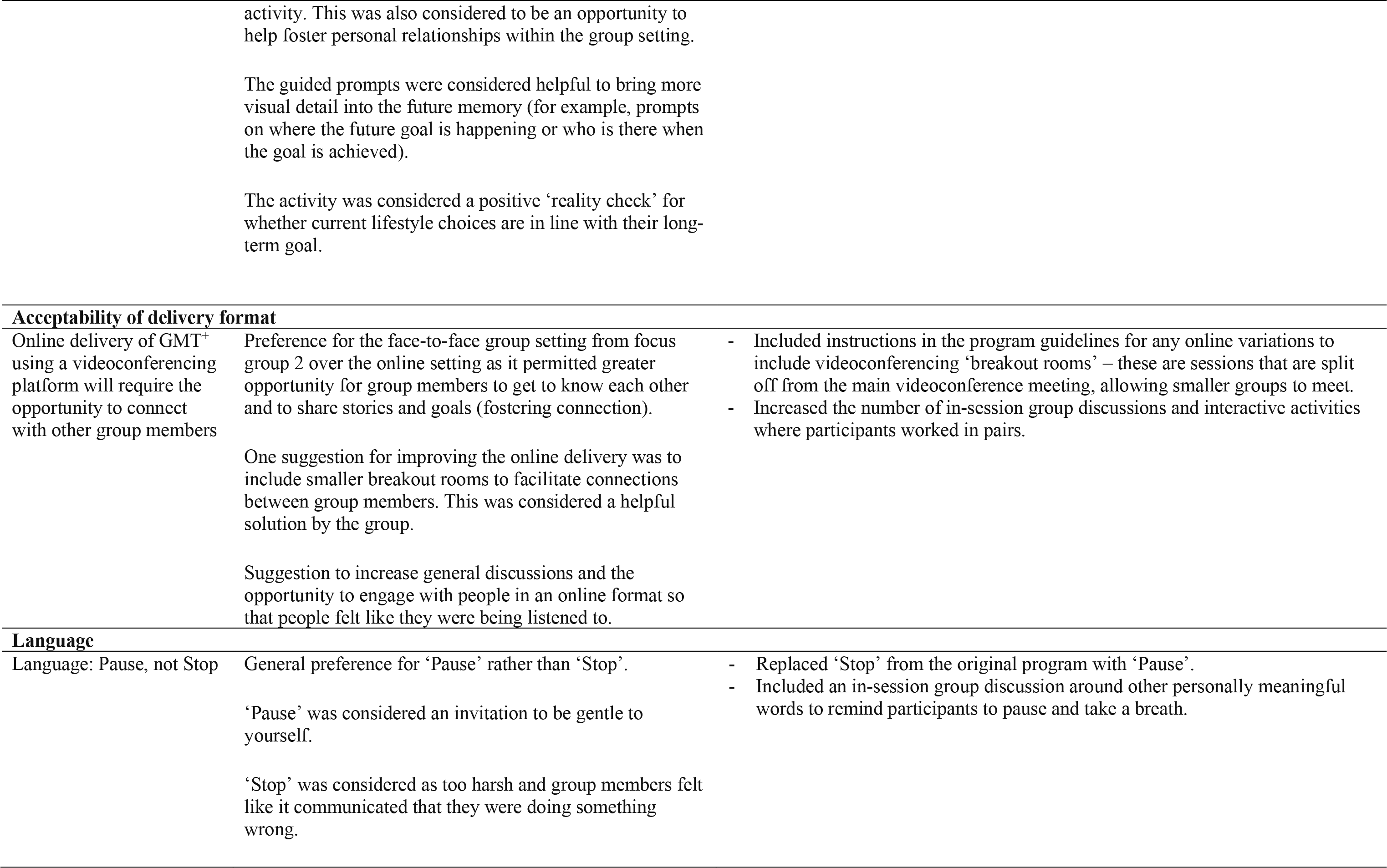
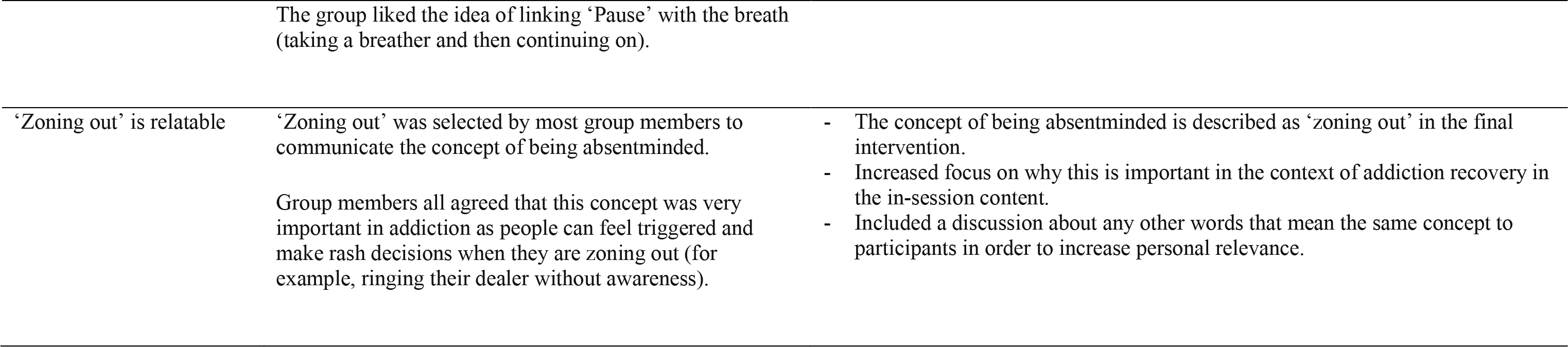
Key feedback from the development (prototype) focus group with people with MUD and subsequent changes to the intervention.

**Figure 7.**
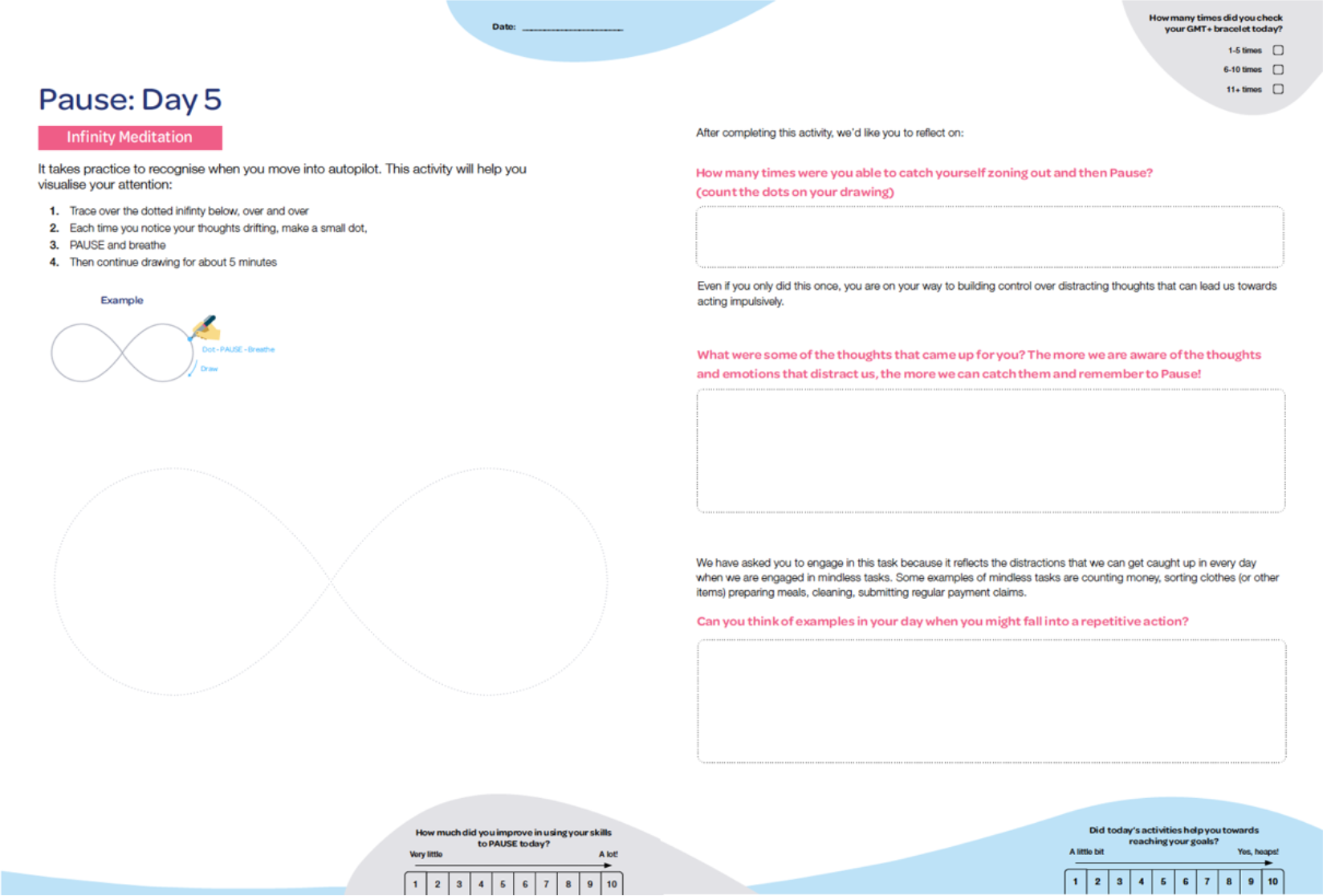
A creative journal activity and guided questions to connect the relevance to everyday life.

Of the five participants who attending the focus group, two completed the sample journal and mailed it back, one participant did not provide their postal address, one indicated forgetting to complete and return the journal, and one reported relocating home during the journal completion period and was unable to mail it back in time. Follow-up phone calls with the two participants who did not mail back the sample journal indicated that they enjoyed engaging with the program and were implementing GMT^+^ skills after the focus group.

#### Results

##### Focus group 3: consumers with MUD (teleconferencing session)

Participants who had attended focus group 2 were positive about how we had incorporated their previous feedback and expressed enthusiasm for the redesigned materials. Sharing how participants’ feedback was implemented is important for building trust in and establishing commitment to the participatory design approach^26^. Feedback from focus group 3 indicated overall acceptability of the language used, the interactive group intervention format, and the content that was tailored to the needs of people with MUD. However, as indicated in Table 4, a key theme that emerged from the think aloud strategy was the need to further enhance the relevance of the program to people who are undergoing treatment for MUD. This included a greater focus on group discussions and characters with more relevant problems for this population. We responded to this feedback by addressing substance use more frequently in group discussions, developing a range of GMT^+^ characters with different demographics, attributes, and goals, and inviting consumers to select their preferred GMT^+^ ambassador character to relate to in-session.

Participants indicated acceptability of the journal and affirmed that they would complete it if enrolled in the GMT^+^ program. Both participants indicated that they felt confident completing the journal activities based on the in-session strategies, discussions, and instructions provided. Finally, participants felt that the types of activities and the language used were appropriate. Participants did not complete the journal daily and one participant stated that they would be more likely to complete it regularly in an inpatient treatment setting. Completion rates and feedback highlighted the importance of reviewing journal activities at the beginning and end of each session to enhance motivation and discuss individual experiences.

We received important feedback on ways to improve the journal. Responses indicated that although the updated single-focussed creative activities were endorsed, (e.g., drawing a repetitive pattern to invoke autopilot) they required greater explanation about the relevance to GMT^+^ skills and everyday life. Participants enjoyed reflecting on their journal work and often provided written content beyond the provided space, suggesting a need for more reflection space on relevant pages. We addressed this feedback by providing a debrief page after each creative activity that explained the relevance of the activity and allowed consumers to reflect on how it may relate to their own life. We increased the reflection text entry space across all activities.

### Phase 4: Intervention Development - Clinical Acceptability

#### Aim

To manualise the intervention (i.e., prepare the presentation slides and journal for administration) and showcase it to clinicians for final feedback prior to implementation.

#### Methods

Clinical psychology researchers and design researchers in healthcare collaborated in developing journal activities that appeared ‘enjoyable’ to complete, whilst training the relevant cognitive skills. The journal activities for each week targeted building attention and meta-cognition (week 1: Be Aware), learning to pause and gain control over impulsivity (week 2: Pause), improving focus on current and short-term goals (week 3: Envision Goals), and improving future-focussed reflection and long-term decision-making skills (week 4: Decide). Once the materials were complete, we conducted a review session with clinical treatment providers to assess reactions to the program and to further optimise acceptability and feasibility.

##### Review session with clinicians

We conducted a 2-hour online review session to present the final program and receive feedback from clinicians (n = 2). Clinicians were recruited from Turning Point and The Turner Clinics, Monash University. We presented the full program including: the in-session presentation slide material and activities, and the between-session journal activities for each of the four modules. Following the review session, we incorporated final clinical feedback into the program materials.

## Results

### Review session with clinicians

Clinical treatment providers in the final review session were positive about the need for this type of intervention in an addiction treatment services context and considered the intervention feasible to implement. Specifically, one clinician commented favourably about how theory has been interwoven into practice “to make the whole package usable for the end-consumer.” The activities were also considered “fresh” and engaging, and appropriate for addiction treatment as the content of the stimuli is far-removed from drug-related stimuli (e.g., replacing playing cards with cartoon and travel post cards). Session feedback is outlined in Table 5. Key feedback included the need to normalise errors and slips to consumers who may be particularly sensitive to making mistakes. We addressed this feedback by highlighting how these errors are experienced by all people and provided relatable examples of mistakes by facilitators. Clinicians also highlighted that one task which promotes multi-tasking (even if tasks were not completed) may foster complacency with goals and reinforce feelings of hopelessness. We included a debrief script to reinforce the objective of making progress towards multiple goals at the same time (to avoid neglecting one goal in favour of another).

**Table 5.** Key feedback from clinical service providers and subsequent changes to the final intervention.

| **Theme** | **Feedback** | **Program modification** |
| --- | --- | --- |
| Normalising errors on the cognitive tasks | People with addiction are hard on themselves when they make a mistake.<br>It is important to normalise that all people make these mistakes. | - Increased focus on normalising errors and communicating that many people regularly make these types of slips.<br>- Explained why it is important to focus on in the context of addiction recovery but used modified language to soften the message. |
| Important to help participants manage multiple long-term goals | People in addiction treatment can become overly focussed on one or two of the goals and end up neglecting the other goals.<br>Consider how you can help participants achieve one of their goals without neglecting the other goals. | - Introduced a brief activity in module 4 (Decide) to task-split multiple goals so that participants can work towards achieving small steps within multiple goals each day. |
| Multitasking activity may send a confusing message as the goal is to attempt all activities, even if they are not completed within the allocated time | People with MUD may often start multiple tasks without finishing them and may often move between tasks. They may feel as though they're failing because they never complete anything.<br>The key here is to be clear about what the task is about – multitasking. | - Provided clearer instructions prior to commencing the task that the purpose is to practice multitasking.<br>- Debrief afterwards to explain that the goal was not to leave each task incomplete, but to focus on the overall goal when working with multiple tasks or components at once. |

### Key intervention modifications

We have made a number of changes to GMT^+^ throughout the planning, intervention design and development phases. Some of the major changes included: 1) developing more relevant and meaningful program characters and narratives to help demonstrate everyday problems where GMT^+^ may help; 2) adapting the language to more appropriately connect the concepts to everyday life for people with MUD; 3) developing more engaging and cognitively appropriate in-session activities; 4) developing a completely new between-session journal to increase the ‘hands-on’ enjoyment of regular skills practice; and 5) including new strategies to develop a future focussed mindset to help with decision making. We describe the final GMT^+^ intervention, guided by Items 3-10 from the TIDieR checklist.

### Goal Management Training^+^

Goal Management Training^+^ (GMT^+^) is a manualised therapist-facilitated group program. We developed a final kit comprising a range of materials to administer GMT^+^. The in-session materials include presentation slide content with manualised presenter scripts for each of the four modules, guided discussions, audio recordings of meditation scripts, and cognitive activity materials (e.g., buzzers, sorting cards). The between-session material includes a printed journal to distribute to each participant, consisting of daily activities that relate to the weekly module (i.e., Be Aware, Pause, Envision Goals, Decide) for them to complete. Sample GMT^+^ materials can be accessed by emailing the corresponding authors. We have included a breakdown of content, in-session activities, and between-session journal activities in Table 6.

**Table 6.**
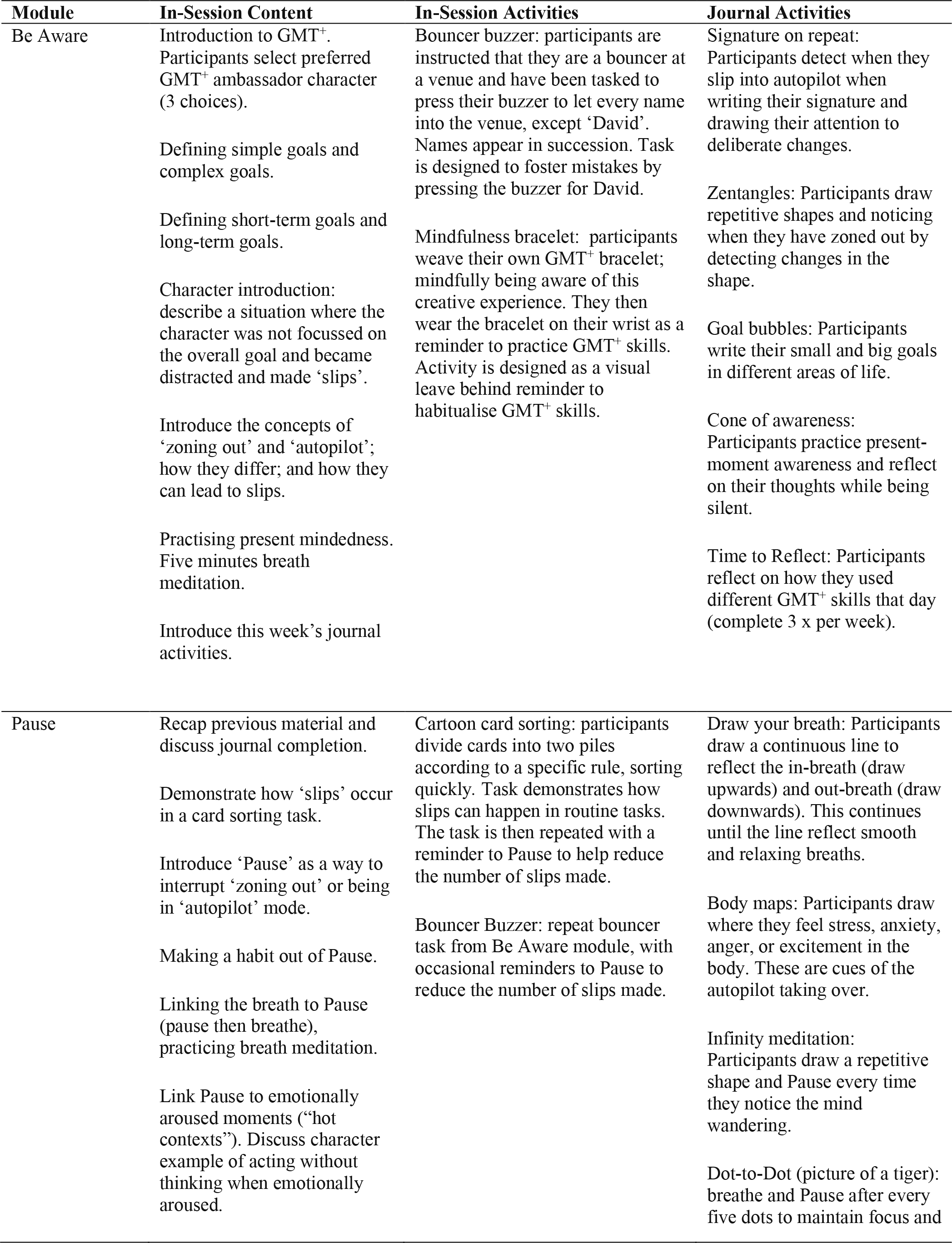
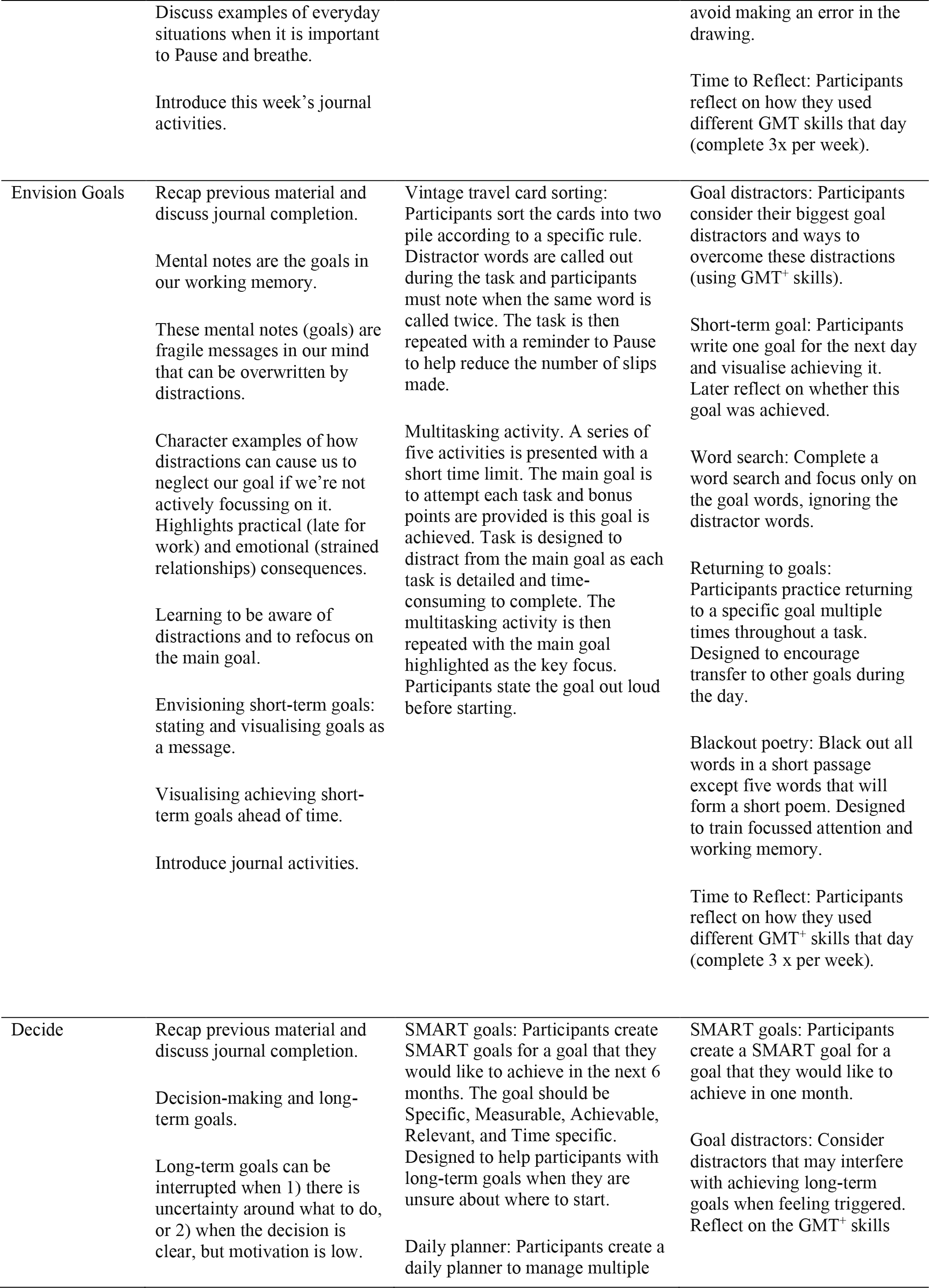
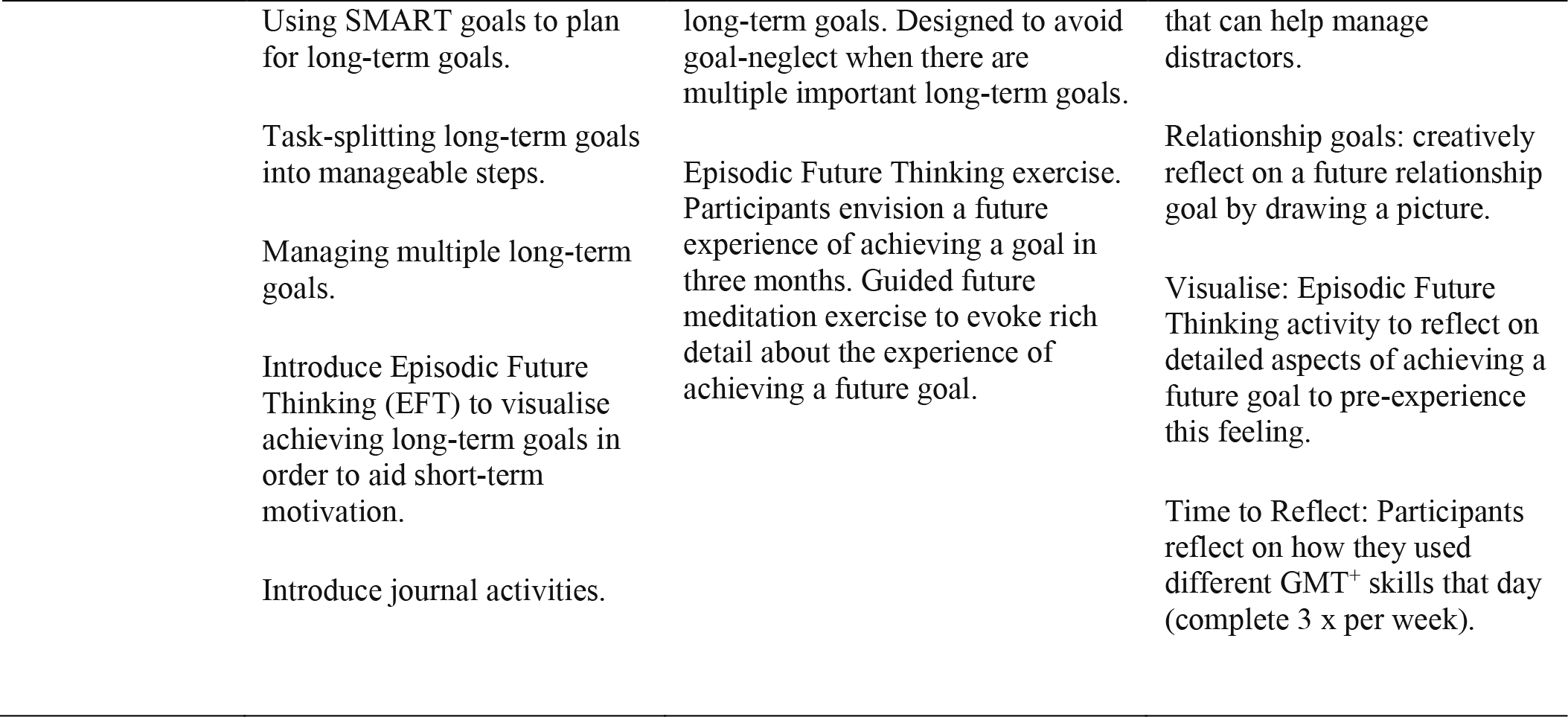
Description of in-session content, in-session activities and between-session journal activities for the four GMT^+^ modules

| Module | In-Session Content | In-Session Activities | Journal Activities |
| --- | --- | --- | --- |
| Be Aware | Introduction to GMT<sup>+</sup>. Participants select preferred GMT<sup>+</sup> ambassador character (3 choices).<br>Defining simple goals and complex goals.<br>Defining short-term goals and long-term goals.<br>Character introduction: describe a situation where the character was not focussed on the overall goal and became distracted and made ‘slips’.<br>Introduce the concepts of ‘zoning out’ and ‘autopilot’; how they differ; and how they can lead to slips.<br>Practising present mindedness. Five minutes breath meditation.<br>Introduce this week’s journal activities. | Bouncer buzzer: participants are instructed that they are a bouncer at a venue and have been tasked to press their buzzer to let every name into the venue, except ‘David’. Names appear in succession. Task is designed to foster mistakes by pressing the buzzer for David.<br>Mindfulness bracelet: participants weave their own GMT<sup>+</sup> bracelet; mindfully being aware of this creative experience. They then wear the bracelet on their wrist as a reminder to practice GMT<sup>+</sup> skills. Activity is designed as a visual leave behind reminder to habitualise GMT<sup>+</sup> skills. | Signature on repeat: Participants detect when they slip into autopilot when writing their signature and drawing their attention to deliberate changes.<br>Zentangles: Participants draw repetitive shapes and noticing when they have zoned out by detecting changes in the shape.<br>Goal bubbles: Participants write their small and big goals in different areas of life.<br>Cone of awareness: Participants practice present-moment awareness and reflect on their thoughts while being silent.<br>Time to Reflect: Participants reflect on how they used different GMT<sup>+</sup> skills that day (complete 3 x per week). |
| Pause | Recap previous material and discuss journal completion.<br>Demonstrate how ‘slips’ occur in a card sorting task.<br>Introduce ‘Pause’ as a way to interrupt ‘zoning out’ or being in ‘autopilot’ mode.<br>Making a habit out of Pause.<br>Linking the breath to Pause (pause then breathe), practicing breath meditation.<br>Link Pause to emotionally aroused moments (“hot contexts”). Discuss character example of acting without thinking when emotionally aroused. | Cartoon card sorting: participants divide cards into two piles according to a specific rule, sorting quickly. Task demonstrates how slips can happen in routine tasks. The task is then repeated with a reminder to Pause to help reduce the number of slips made.<br>Bouncer Buzzer: repeat bouncer task from Be Aware module, with occasional reminders to Pause to reduce the number of slips made. | Draw your breath: Participants draw a continuous line to reflect the in-breath (draw upwards) and out-breath (draw downwards). This continues until the line reflect smooth and relaxing breaths.<br>Body maps: Participants draw where they feel stress, anxiety, anger, or excitement in the body. These are cues of the autopilot taking over.<br>Infinity meditation: Participants draw a repetitive shape and Pause every time they notice the mind wandering.<br>Dot-to-Dot (picture of a tiger): breathe and Pause after every five dots to maintain focus and |
|  | Discuss examples of everyday situations when it is important to Pause and breathe. |  | avoid making an error in the drawing. |
|  | Introduce this week's journal activities. |  | Time to Reflect: Participants reflect on how they used different GMT skills that day (complete 3x per week). |
| Envision Goals | Recap previous material and discuss journal completion.<br>Mental notes are the goals in our working memory.<br>These mental notes (goals) are fragile messages in our mind that can be overwritten by distractions.<br>Character examples of how distractions can cause us to neglect our goal if we're not actively focussing on it. Highlights practical (late for work) and emotional (strained relationships) consequences.<br>Learning to be aware of distractions and to refocus on the main goal.<br>Envisioning short-term goals: stating and visualising goals as a message.<br>Visualising achieving short-term goals ahead of time.<br>Introduce journal activities. | Vintage travel card sorting: Participants sort the cards into two pile according to a specific rule. Distractor words are called out during the task and participants must note when the same word is called twice. The task is then repeated with a reminder to Pause to help reduce the number of slips made.<br>Multitasking activity. A series of five activities is presented with a short time limit. The main goal is to attempt each task and bonus points are provided if this goal is achieved. Task is designed to distract from the main goal as each task is detailed and time-consuming to complete. The multitasking activity is then repeated with the main goal highlighted as the key focus. Participants state the goal out loud before starting. | Goal distractors: Participants consider their biggest goal distractors and ways to overcome these distractions (using GMT<sup>+</sup> skills).<br>Short-term goal: Participants write one goal for the next day and visualise achieving it. Later reflect on whether this goal was achieved.<br>Word search: Complete a word search and focus only on the goal words, ignoring the distractor words.<br>Returning to goals: Participants practice returning to a specific goal multiple times throughout a task. Designed to encourage transfer to other goals during the day.<br>Blackout poetry: Black out all words in a short passage except five words that will form a short poem. Designed to train focussed attention and working memory.<br>Time to Reflect: Participants reflect on how they used different GMT<sup>+</sup> skills that day (complete 3 x per week). |
| Decide | Recap previous material and discuss journal completion.<br>Decision-making and long-term goals.<br>Long-term goals can be interrupted when 1) there is uncertainty around what to do, or 2) when the decision is clear, but motivation is low. | SMART goals: Participants create SMART goals for a goal that they would like to achieve in the next 6 months. The goal should be Specific, Measurable, Achievable, Relevant, and Time specific. Designed to help participants with long-term goals when they are unsure about where to start.<br>Daily planner: Participants create a daily planner to manage multiple | SMART goals: Participants create a SMART goal for a goal that they would like to achieve in one month.<br>Goal distractors: Consider distractors that may interfere with achieving long-term goals when feeling triggered. Reflect on the GMT<sup>+</sup> skills |

|  |  |  |
| --- | --- | --- |
| Using SMART goals to plan for long-term goals. | long-term goals. Designed to avoid goal-neglect when there are multiple important long-term goals. | that can help manage distractors. |
| Task-splitting long-term goals into manageable steps. | Episodic Future Thinking exercise. Participants envision a future experience of achieving a goal in three months. Guided future meditation exercise to evoke rich detail about the experience of achieving a future goal. | Relationship goals: creatively reflect on a future relationship goal by drawing a picture. |
| Managing multiple long-term goals. |  | Visualise: Episodic Future Thinking activity to reflect on detailed aspects of achieving a future goal to pre-experience this feeling. |
| Introduce Episodic Future Thinking (EFT) to visualise achieving long-term goals in order to aid short-term motivation. |  | Time to Reflect: Participants reflect on how they used different GMT<sup>+</sup> skills that day (complete 3 x per week). |
| Introduce journal activities. |  |  |

GMT^+^ facilitators should be familiar with the content and manuals prior to administering the intervention. The current intervention is designed as an in-person (face-to-face) intervention that should be delivered on-site at an inpatient addiction treatment facility. The facilitator will need access to a projector screen and computer (to run the presentation slide content), tables (as participants work with physical materials) and a quiet room. There are four weekly sessions that run for 90 minutes and should be delivered in groups of 4-8 program participants. The journal contains detailed instructions for daily completion and does not require any input or participant monitoring from staff at the treatment facility. Between-session journal completion is discussed as a group at the beginning of sessions 2-4 and the facilitator will assess for daily completion.

## Discussion

This study aimed to update GMT (a cognitive remediation program for brain injury) to tailor it to the key cognitive deficits and treatment context of MUD, and to maximise users’ engagement with its contents and delivery. We utilised an evidence- and person-based approach, collaborating with clinical stakeholders, neuropsychology experts, designers in healthcare, and people with MUD to develop the four-week GMT^+^ program. Results from our four-phased approach revealed that GMT^+^ can be feasibly and ecologically applied in addiction treatment settings and has shown initial evidence of consumer engagement. These findings illustrate the benefits of the evidence- and person-based approach and provide confidence to move into the evaluation phase with a protocol that minimises potential risks, for example, barriers to implementation, or a lack of consumer engagement.

The initial focus on four cognitive processes (i.e., attention, impulse control, goal setting, decision-making) was endorsed by experts and consumers. We selected these four components based on emerging research revealing impairments in these specific executive functions, as well as their relationship with key addiction treatment outcomes^8, 9, 11, 32^. One of the primary goals of addiction treatment is to help individuals to develop self-control strategies to manage cravings and emotionally salient situations, skills that inherently rely on these prioritised cognitive processes.

The updated GMT^+^ program trains these components through enhancing meta-cognition (‘thinking about thinking’), fostering skill practice in everyday situations, and reinforcing a simplified cycle to employ in any given moment [Be Aware (of inattention) →Pause (to prevent acting on autopilot)→Envision Goals (to prevent goal distractions)→ Decide (to consider longer-term outcomes)]. Consumers build on these skills as they progress through the program, providing an opportunity for skill acquisition and mastery through self-initiated practice and active reflection of cognitive processes and situational consequences. Unlike some existing interventions that train specific cognitive processes through repetitive task practice, e.g., computerised working memory or inhibitory control training^33, 34^, GMT^+^ teaches consumers to group multiple skills together and apply them in ecologically relevant situations (for example, noticing drug-cravings, taking a depth breath, and bringing attention back towards short-term or long-term goals).

Neuropsychology experts highlighted the need to help participants to habitualise strategies to promote their effectiveness in critical real-world situations. We addressed this suggestion with the inclusion of the GMT^+^ mindfulness bracelet to serve as a specific visual cue to promote a stimulus-response association^35^, for example, noticing the bracelet and applying ‘pause’. As this bracelet is always accessible to consumers, it may enhance the chance of successful habit formation^36^. We also incorporated activities in the take-home journal to encourage consistency of practice and skill mastery^36, 37^. For example, a checkbox to note how many times consumers used their bracelet each day, activities to reflect on how different GMT^+^ skills were employed that day, and creative activities that were superficially enjoyable (e.g., dot-to-dot, mindful drawing) but required the use of GMT^+^ skills to complete.

Consumers provided input on their personal goals during recovery and emphasised the need to develop relatable characters and scenarios. We used this feedback to create a range of character profiles and situations where a character may have benefited from applying GMT^+^ strategies. We provided consumers with the opportunity to personalise their group treatment by selecting their preferred GMT^+^ ambassador character to enhance meaning and motivation to engage in group discussions^38^. Consumers indicated a preference for simplified creative journal activities with a clear purpose. We responded to this feedback by creating enjoyable activities requiring brief instructions and including clear links to everyday challenges for consumers to reflect on. We reason that these activities will be well suited to promote strategy habituation due to anticipated adherence to regular journal completion (skills practice). Consumers also preferenced the word ‘Pause’ over ‘Stop’, which was considered punitive. Amending the language and visual representation of key strategies is critical to fostering program engagement and acceptability of the strategies. An unintended benefit might also be to promote self-compassion during a fragile treatment period where self-criticism may be more likely to lead to earlier program drop-outs and re-engagement in addictive behaviours^39, 40^.

The final showcase to clinicians identified important unintended effects, for example, GMT^+^ uses multitasking (even if all activities are not completed) and task errors as ways to illustrate challenges for executive function; however, these aspects may trigger procrastination and perceived hopelessness. We addressed this feedback by enhancing the context around these activities (i.e., explaining the aim as ‘making progress towards goals’). Facilitators are also prompted to share personal accounts of everyday ‘slips’ to group members and to normalise mistakes on the tasks as part of the learning process, with the goal of reducing feelings of shame or a tendency to overidentify with making errors.

### Strengths and limitations

There were several strengths to our design process. We employed four diverse groups in the development process, including neuropsychology experts, design researchers in healthcare, consumers with MUD, and clinical service providers. We conducted both face-to-face and online focus groups with consumers, providing an indication of positive engagement with future online administration. Our person-based, participatory design perspective valued and incorporated the needs and experiences of the end-consumer at each intervention development phase.

There were some limitations to our work. There were only four participants in focus group 2 and five participants in focus group 3, potentially limiting the perspectives provided from consumers with MUD. We included participants with both current and past methamphetamine use. It is possible that the perspectives of those who no longer used methamphetamine are not representative of a population currently undergoing treatment. However, this concern is balanced by the benefit of receiving perspectives from people who understand the treatment process and have maintained abstinence. Further, we only tested a prototype of module 1 (Be Aware) due to time constraints. Although we tested key concepts from other modules (e.g., Pause, the mental notepad, EFT), the presentation slide materials and journal activities for modules 2, 3 and 4 were not delivered in their final form to the end-consumer. However, clinical service providers were positive about the final materials for all four modules.

### Future Research

The main components of GMT^+^ are relevant not only to MUD but also other substance use and addictive disorders. This research may inform the development of modified interventions for different addictions, incorporating similar evidence- and person-based design principles. This paper also highlights the importance of collaborating with end-consumers prior to administering existing evidence-based interventions in different consumer populations. We employed several changes to design, training concepts, relevance of characters, and how the strategies were practiced between sessions to aid habituation.

Our research has provided preliminary support for an online application of GMT^+^, based on the online delivery of the prototype and verbal feedback provided by focus group participants. The cognitive activities require conversion to an online computerised format and additional opportunities for online participant interaction should be considered to maintain engagement. There is also a potential to expand this intervention to treat other mental health needs where there are executive dysfunctions and difficulties with goal-related decisions, for example, binge eating disorder, ADHD, or schizophrenia^14, 41, 42^. This program may also be appropriate to apply to other populations where shorter administration times and less content repetition are indicated (e.g., OCD)^43^. To adapt the intervention for these groups, content changes would need to apply, such as tailoring character examples to these groups and raising discussions about how GMT^+^ skills could be applied for everyday difficulties. Consultation with specific service providers and relevant consumer groups could assist with these changes.

## Conclusions

We have presented the systematic development of GMT^+^, an updated version of GMT that is tailored to key cognitive deficits and treatment requirements for MUD. GMT^+^ is a four-week group program with a between-session journal to foster everyday skill practice. It includes clear and practical strategies to employ in everyday situations and is designed to improve attention, impulse control, goal setting and decision-making in MUD. By employing an evidence- and person-based approach, we have demonstrated how potential barriers to engagement and uptake by consumers were able to be addressed through modifications to the intervention content, materials, and delivery format. We are confident that we have developed an intervention with initial acceptability for the treatment of MUD. Further research is now required to assess the feasibility and efficacy of GMT^+^ in a pilot trial (specific details are included in the trial registry; Trial ID: ACTRN12621000172808). This will determine whether GMT^+^ is effective at improving executive functions and clinical outcomes for patients with MUD and whether it may have utility as an adjunct inpatient addiction treatment.

## Data Availability

The datasets used during the current study are available from the corresponding author on reasonable request.

## Abbreviations

MUD: Methamphetamine Use Disorder
GMT: Goal Management Training
GMT^+^: Goal Management Training^+^

## Ethics approval and consent to participate

Ethical approval for this study was obtained from Monash University Human Research Ethics Committee (12364) and Eastern Health Human Research Ethics Committee (LR19/023). Participants provided informed consent to participate in the focus groups.

## Consent for publication

Not applicable.

## Authors’ contributions

AA and AVG conceived the study. AA, AR, and AVG conducted focus group 1 and interpreted the expert feedback. AA, AR, EP, and BK conducted focus group 2 with consumers and interpreted collective feedback. AA, AR, EP, and BK conducted focus group 3 with consumers and interpreted collective feedback. DF oversaw the research design process. AA and AR presented the final materials to clinical service providers. AVG and DL supervised the study. AA wrote the first draft of the manuscript. All authors contributed to the writing process and approved the final version of the manuscript.

## Acknowledgements

Not applicable.

